# Strengthening diagnostic capacity for viral hemorrhagic fevers in Forest Guinea: advances in Lassa fever case detection

**DOI:** 10.64898/2026.02.24.26346968

**Authors:** Fara Raymond Koundouno, Youssouf Sidibe, Saa Lucien Millimono, Kékoura Ifono, Julia Hinzmann, Hugo Soubrier, Karifa Kourouma, Tamba Elie Millimouno, Fernand M’Bemba Tolno, Faya Moriba Kamano, Mamadou Dioulde Barry, Soua Koulemou, Bely Sonomy, Mariame Traore, Kaba Keïta, Mette Hinrichs, Sarah Ryter, Carolina van Gelder, Beate Becker-Ziaja, Christine Jacobsen, Anke Thielebein, Lisa Oestereich, Meike Pahlmann, Anaïs Legand, Pierre Formenty, Aly Antoine Kamano, Seydou Dia, Moke Fundji Jean Marie Kipela, Stephan Günther, Kolié Moussa, Kaba Keïta, Sira Hélène Guilavogui, Sanaba Boumbaly, N’Faly Magassouba, Sophie Duraffour, Giuditta Annibaldis

## Abstract

Viral hemorrhagic fevers (VHFs), including filoviruses, Lassa virus, yellow fever virus and dengue virus, remain a major public health threat in resource-limited settings. The 2014–2016 Ebola epidemic exposed critical gaps in laboratory preparedness and diagnostic capacity in most-at-risk countries, including Guinea. In order to implement sustainable in-country laboratory capacity for early detection and rapid response to VHF outbreaks, a decentralized diagnostics laboratory strengthening program started in the forested region of Guinea in Guéckédou (2017) and N’Zérékoré (2021).

Between 2017 and 2024, a total of 4683 samples from suspected VHF cases were tested across the two laboratories. Of those, 47 (1%) were identified as positive for VHFs by molecular testing, including the 2021 Ebola virus disease resurgence and Marburg virus disease emergence samples, for which the laboratories supported rapid outbreak detection and case management. Notably, between 2020 and 2024, 30 Lassa fever cases were laboratory-confirmed with a case fatality rate of 67.9%. Of those, 66.7% were men, with 50.0% and 27.6% of the cases originating from the Guéckédou and N’Zérékoré prefecture, respectively. Prior to this program, Lassa fever detection in Guinea remained anecdotic. One case of yellow fever and one of dengue were also identified by molecular assays.

The establishment of locally embedded laboratory infrastructure enhanced VHFs preparedness, surveillance, and response in Guinea. Beyond improving understanding of Lassa fever epidemiology, this program highlights the value of sustained laboratory capacity for early detection and health system resilience in VHF-endemic settings.

## Introduction

Since the adoption of the International Health Regulations in 2005, the World Health Organization (WHO) has declared eight Public Health Emergencies of International Concern, five of which originated in low-resource settings. Laboratory infrastructure and trained workforce, where needed, allows for timely detection and response to such infectious diseases related emergencies. Of those, viral hemorrhagic fevers (VHFs), including Ebola disease and Lassa fever (LF), pose a major public health threat [1].

The 2014–2016 Ebola virus disease (EVD) epidemic in West Africa exposed critical weaknesses in healthcare systems of affected countries with, among others, the limited testing capacity for VHFs and local trained workforce [2,3]. Particularly in Guinea, the epicenter of the 2014-2016 EVD epidemic, this limited capacity contributed to the delayed detection of the first Ebola case in the forested region, which in turn postponed outbreak confirmation and timely implementation of control measures [4]. Overall outbreak control efforts, given the unprecedented dimension of the epidemic and scarce in-country capacities, relied on the deployment of mobile laboratories staffed with international experts [5–7]. Among those, the European Mobile Laboratory (EMLab, Germany) started diagnostics operations in Guéckédou (Guinea) as early as March 2014 under the umbrella of the Ministry of Health (MoH) and the WHO [6]. Along two years, the EMLab, together with local resources integrated in its daily laboratory operations, pursued its diagnostics mandate, relocating in function of the epidemic dynamics. Preposing decentralized laboratory capacities became an incredible asset in controlling the outbreak and detecting EVD flare-ups [8]. As the epidemic was waning, enabling the continuation of laboratory surveillance activities in remote locations (i.e., distant form the reference laboratory in the capital town) to mitigate future public health emergencies became Guinea’s MoH priority [8].

To this end, a legacy program for laboratory strengthening in VHF-prone areas was initiated in 2016 together with the VHFs reference laboratory in Conakry (*Centre de Recherche en Virologie* – *Laboratoire des Fièvres Hémorragiques de Guiné*e, CRV-LFHVG), the *Agence Nationale de Sécurité Sanitaire* (ANSS) of Guinea’s MoH, the Guéckédou prefectural hospital, and the *Direction Préfectorale de la Santé* (DPS). As result, one laboratory was established in Guéckédou at the end of 2016, and a second strengthened in N’Zérékoré in 2021, providing support to the national surveillance system with molecular diagnostics for VHFs at no expense.

VHFs are caused by zoonotic RNA viruses maintained in animal reservoirs such as rodents, bats or arthropods, with transmission occurring through contact with infected hosts or via human-to-human spread [9]. Following variable incubation periods, they typically present with non-specific symptoms (including fever, malaise, headache and myalgia) that may progress to severe disease characterized by hemorrhage, shock and multi-organ failure. Clinical diagnosis remains challenging due to overlapping presentations with other infections, and confirmation relies on molecular laboratory assays, mostly real-time reverse transcription polymerase chain reaction (RT-PCR). Management is primarily supportive, as therapeutic options are limited, with ribavirin used for Lassa virus (LASV) infections although evidence for its efficacy is anecdotal [10]. Licensed vaccines and or other countermeasures are available only for a subset of these diseases; however, access and coverage remain constrained in many endemic settings [9,11]. In Guinea, beyond the EVD outbreak, acute LF cases have been rarely reported [12–15] despite serological evidence of widespread population exposure in the central and forest regions [16–18]. Yellow fever is endemic in the country, with sporadic outbreaks linked to sylvatic transmission and low vaccination coverage [19], while dengue fever is still underreported due to surveillance system limitation. The presence of established wildlife reservoirs in the Forest region, including rodents for LASV [20] and bats for filoviruses (i.e., *Orthomarburgvirus marburgense* and *Orthoebolavirus genus*) [21,22], support sustained zoonotic spillover risks and underline the epidemic potential in the area. Prior the establishment of the two laboratories in Guéckédou and N’Zérékoré, detection of VHFs cases in country relied on the national reference laboratory in Conakry, restricting the surveillance, particularly in the forested area, and leading to scarce published data on VHF surveillance and epidemiology in Guinea.

Here, we report on VHFs diagnostics surveillance activities, including LASV monitoring, conducted by two decentralized laboratories located in Forest Guinea, highlighting laboratory capacity strengthening, epidemic preparedness activities, and support to public health response.

## Methods

### Ethics

This research has been approved by the National Ethics Committee of Guinea (CNERS) under the numbers 197/CNERS/25 and 189/CNERS/24.

### Samples, case definition and case management

Samples (whole blood EDTA or buccal swabs) from suspected VHF cases were collected by the national routine surveillance teams and or clinical teams and dispatched for testing to the *Laboratoire des Fièvres Hémorragiques Virales de Guéckédou* (LFHV-GKD), Guéckédou, or *Laboratoire des Fièvres Hémorragiques Virales de l’Hôpital Régional de N’Zérékoré* (LFHV-HRNZE), N’Zérékoré. LFHV-GKD was operational in 2017 and LFHV-HRNZE in 2021. Suspected and confirmed VHF cases were classified in accordance to the Guinea’s national guidance document «*Guide technique national de surveillance intégrée de la maladie et de la riposte (SIMR), Guinée (2019)*» [23]. A suspected case was defined as any critically ill patient or deceased individual with an acute onset of fever together with hemorrhagic manifestation(s) or where there was clinical suspicion of viral infection. A confirmed case was defined as a suspected case with laboratory confirmation by a positive real-time RT-PCR result for the pathogen of interest. Confirmed patients were managed in designated Treatment Centre for Epidemics (CTEPi; *Centre de Traitement des Épidémies*) and clinical care provided according to national protocols. If available, Lassa fever patients received ribavirin according to the McCormick regimen [24]. Patients were discharged after clinical recovery, with RT-PCR confirmation of viral clearance performed at the discretion of the physician.

### Ecological investigations at LFHV-GKD

Investigations on small mammals were coordinated by the Ministry of the Environment (*Ministère de l’Environnement et du Développement Durable (MEDD)*) in the context of an outbreak response. Rodents were randomly captured following the methodology as described in [25]. Autopsies were performed in the field with blood, liver, spleen, and feces collected and further stored at −20 °C until laboratory processing. Organs were homogenized and stored in phosphate buffer saline (PBS). Samples were pooled and processed for viral RNA extraction real time RT-PCR for Lassa virus as described in the laboratory procedures section. Positive pools were identified and individual samples subsequently tested by RT-PCR.

### Laboratory procedures

Samples were inactivated as previously described [26]. Briefly, viral RNA was extracted with the QIAamp viral RNA extraction kit (Qiagen, Germany) according to manufacturer’s instructions modified with two rounds of buffer AW2 washes and a 10-minute dry spin before elution. RNA extracts were used for RT-PCR on the Rotor-Gene Q platform (Qiagen); leftover RNA was stored at −20°C. The RT-PCR assays varied based on the year and sites as follows. Note that all kits named after RealStar^®^ are from the company altona Diagnostics (Germany). At LFHV-GKD, filoviruses (enabling distinct detection between *Orthomarburgvirus marburgense* (for ease of reading, abbreviated here as MARV) and *Orthoebolavirus* (for ease of reading, abbreviated here as EBOV) virus species) were tested using the RealStar^®^ Filovirus Screen RT-PCR Kit 1.0. LASV diagnostics from 2017 to 2019 was done with RealStar^®^ Lassa Virus RT-PCR Kit 1.0 targeting the S segment, and an in-house version of the Nikisins RT-PCR targeting the L-segment as described in [27]. Since 2019, the RealStar^®^ Lassa Virus RT-PCR Kit 2.0, targeting both the S and L segments, was in use. For Yellow fever virus (YFV), RT-PCR assays included that of Domingo et al. [28] and the RealStar^®^ Yellow Fever Virus RT-PCR Kit 1.0. For dengue virus (DENV), the RealStar^®^ Dengue Virus RT-PCR Kit 3.0 was used. At LFHV-HRNZE, filoviruses were tested using the RealStar^®^ Filovirus Screen RT-PCR Kit 1.0. The testing of EBOV was also done using the Xpert^®^ Ebola (GeneXpert^®^, Cepheid^®^, U.S.). Additionally, the OraQuick™ Ebola Rapid Antigen Test (OraSure Technologies Inc., U.S.) was also used for screening community or hospital deaths (oral swabs) during EVD in 2021. The following RT-PCR assays were used for the pathogen(s) of interest, RealStar^®^ Lassa Virus RT-PCR Kit 1.0, RealStar^®^ Yellow Fever Virus RT-PCR Kit 1.0, and RealStar^®^ Dengue Virus RT-PCR Kit 3.0.

LASV-positive samples were tested for anti-LASV nucleoprotein (NP) immunoglobulins G and M (IgG and IgM) using the Panadea ELISA kits (Panadea GmbH, Hamburg, Germany) per manufacturer instructions. Serological testing was performed outside the routine diagnostic activities, and only when sufficient sample volume was available.

### Data management and statistical analysis

Both laboratories used similar quality management systems and processes in the laboratory. Basic demographic data, recorded on the laboratory request form and accompanying the samples sent to the laboratory for testing, were entered into the laboratory database (Excel, Microsoft). Laboratory diagnostic findings were then added to the database. For LASV positive cases, where applicable, related clinical (including symptoms at presentation) and or epidemiological and or ecological data were retrospectively extracted from unstructured medical records and linked. For this analysis, the databases were anonymized and all diagnostics tests included in the LFHV-GKD database from January 2017 to December 2024, and that of LFHV-HRNZE from February 2021 to December 2024, were included. The databases were curated prior to analysis excluding follow up tests of confirmed cases performed for patient monitoring and the final statistics for each laboratory only display initial diagnosis results (first test). Statistical analysis was performed using STATA software (v.18, StataCorp, USA). Graphs were generated using GraphPad Prism v.10 Software (USA). Maps were produced using the QGIS Geographic Information System software (version 3.26, Open-Source Geospatial Foundation Project), with administrative boundary data downloaded from the GADM database (v.4.1, Global Administrative Areas).

## Results

### Establishment of laboratory-based VHFs surveillance in Forest Guinea

A long-term laboratory capacity-strengthening program for VHFs preparedness was initiated in late 2016 in Forest Guinea, near the epicenter of the West African EVD outbreak. This led to the establishment of a diagnostic laboratory in Guéckédou, LFHV-GKD, under the umbrella of the national reference laboratory for VHFs in Conakry (CRV-LFHVG) and in collaboration with the Bernhard Nocht Institute for Tropical Medicine (BNITM, Germany). Even if not setup within the hospital due to lack of space, LFHV-GKD was connected to the prefectural hospital of Guéckédou and operated under the mandate of the MoH, and of the ANSS, with support of the DPS. Since its inception, LFHV-GKD was embedded within the national surveillance system for VHFs. Over eight years, the program combined infrastructure reinforcement and workforce development to strengthen national VHF diagnostic and surveillance. Generators and solar energy were setup to enable laboratory activities as public power access remains unavailable in the Guéckédou prefecture. Local staff received intensive hands-on training in biosafety and biosecurity, quality management systems, and molecular diagnostic tests specific for VHFs through expert-led workshops, followed by supervised practice, continuing mentorship, and remote technical support. A train-the-trainer model to sustain competency was setup. Since 2017, LFHV-GKD has provided the Guinea surveillance system with locally accessible, cost-free molecular diagnostic testing for multiple VHFs, including filoviruses (e.g., EBOV and MARV), as well as LASV, YFV, and DENV (**Figure 1; Table S1**).

**Figure 1.**
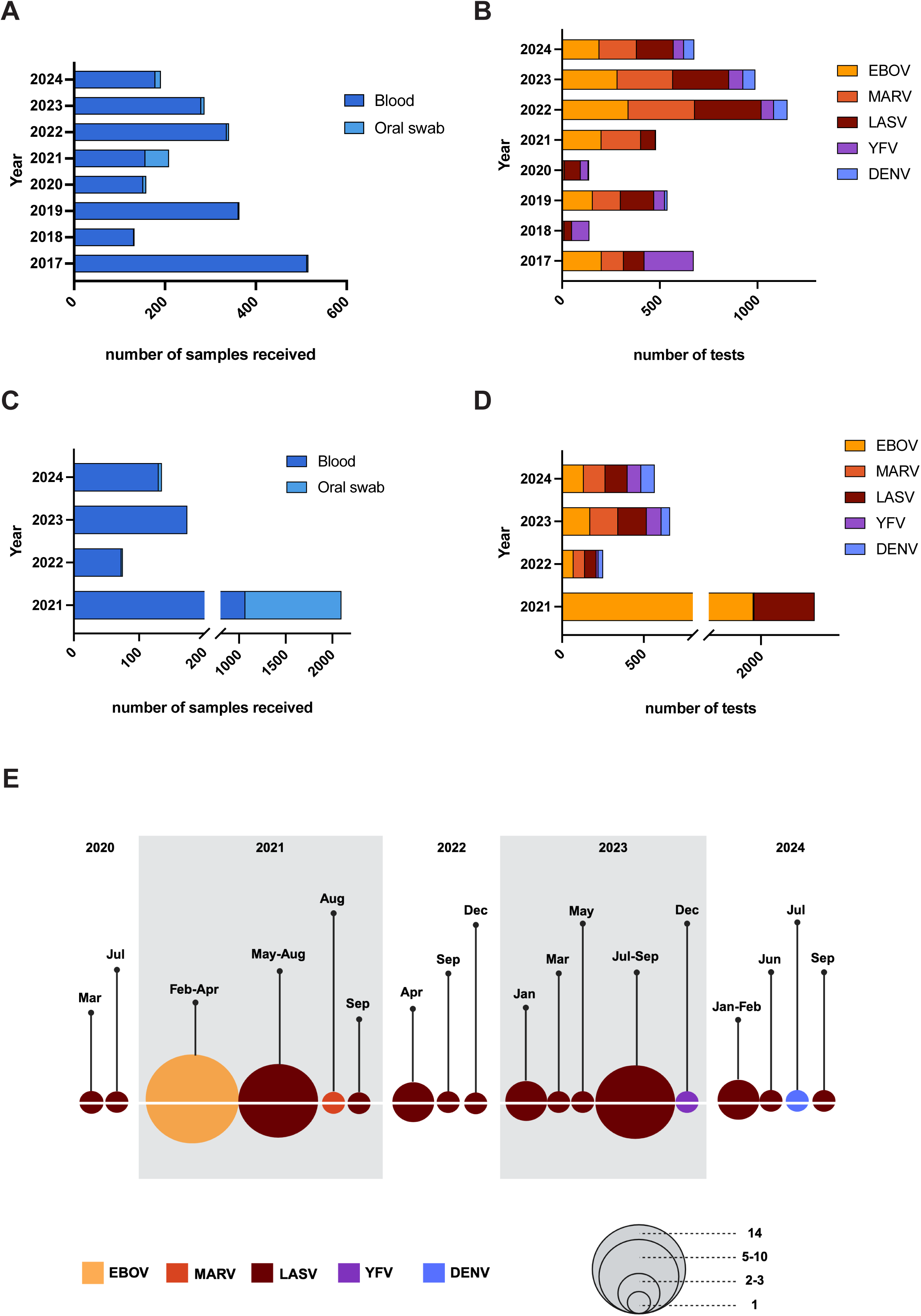
Annual number of samples received, tests performed and positive cases detected for each pathogen at two laboratories. (A) Number of samples received annually, and (B) number of real-time RT-PCR tests conducted per year at LFHV-GKD, Guéckédou. (C) Number of samples received annually, and (D) number of real-time RT-PCR tests conducted per year at LFHV-HRNZE, N’Zérékoré. (E) Number of VHFs positive cases detected over time by the two laboratories (panel created with Biorender). Bubble size represents the number of cases, and the color indicates the pathogen identified. Data are summarized from Tables S1 and S3. Pathogens are indicated as Ebola virus (EBOV), Marburg virus (MARV), Lassa virus (LASV), Yellow fever virus (YFV), dengue virus (DENV).

LFHV-HRNZE, which was initially established in 2014 for the EVD response and survivor program under the umbrella of CRV-LFHVG, joined the VHF capacity development program in 2021 during the resurgence of EVD. Already integrated in the national surveillance system for VHFs, LFHV-HRNZE was at the epicenter of the 2021 resurgence and thus its infrastructure and workforce got reinforced with upgraded laboratory processes and differential VHF testing setup (**Figure 1; Table S1**). Both public power and generators were available to support sustained lab activities as part of the Regional Hospital of N’Zérékoré. Since 2021, laboratory training and technical support were provided in collaboration with and by LFHV-GKD, CRV-LFHVG, and BNITM. Since then, both LFHV-GKD and -HRNZE perform molecular testing for VHFs in a harmonized manner according to similar standard operating procedures (SOPs), which are also implemented in the national reference laboratory CRV-LFHVG.

At start of operations, all laboratory positive diagnostic findings were systematically checked together with, and confirmed by, the national reference laboratory CRV-LFHVG (i.e., sending of an aliquot of the positive sample and or quality assurance management verification) and the BNITM (quality assurance management verification) before official disclosure. Laboratory results were then promptly communicated to the health authorities via official channels in place to ensure public health response and case management at prefectural, regional and country levels.

### Diagnostic activities by the strengthened laboratory capacities

#### LFHV-GKD

Between January 2017 and December 2024, LFHV-GKD tested a total of 2200 samples (2111 EDTA-blood and 89 oral swabs) collected from suspected VHFs cases sent via the national surveillance system (**Table S1**). Sample numbers varied annually, ranging from 133 to 516. The number of tests conducted also fluctuated; for example, in 2022, a total of 1154 real-time RT-PCR assays were performed on 341 samples, corresponding to approximatively 3.4 tests per sample (**Figure 1, A-B; Table S1**). This is reflected in uneven distributions across the years, partly attributable to (i) limited availability of diagnostic kits in the laboratory, as observed in 2018; (ii) reduced diagnostic activity during the COVID-19 pandemic in 2020; and (iii) variations in differential diagnosis driven by clinicians’ requests (**Figure 1, A-B**). Specimens mostly originated from individuals living in Guinea (99.5%, data not shown) and occasionally from travelers from Liberia and Sierra Leone. Samples were substantially skewed towards the prefecture of Guéckédou (95.8%, **Table S2**) where the laboratory is located. Other specimens originated from nearby prefectures, including N’Zérékoré and Yomou, as well as others (**Table S2**). Both sexes were represented equally (56.5% female) and the median age was 35 years [IQR 24-50, min-max: 1-95]. Occupations included domestic workers (34.7%), students (16.2%), farmers (15.3%), health workers (3.4%) and other (27.4%) or unknown (3.1%).

RT-PCR positivity rates for EBOV and MARV were respectively at 4.0% and 0.5% in 2021 (described elsewhere, [26,29]), while undetected in other years (**Figure 1, E**; **Table S1**). LASV showed an RT-PCR positivity rate ranging from 0.6% to 2.8% annually throughout the surveillance period (**Figure 1, E**; **Table S1**). Of the 626 total samples tested for YFV, only one of 72 in 2023 was found positive by RT-PCR (**Figure 1, E**; **Table S1**). More consistent testing for DENV by real-time RT-PCR was setup in 2022 while it was inconsistent in previous years, leading to a total of 201 samples tested (**Table S1**).

From 2017 to 2024, the median turnaround time (TaT) from sample collection to laboratory result was of one day [IQR 0-2, min max: 0-43]. The median time between symptom onset and sample collection was 4 days [IQR 2–8, min-max: 0-184] with still 20% of the overall sample set harboring a time > 10 days (data not shown). Notably, 45% of the samples received in 2017 had a time from onset to sample collection > 10 days which dropped to 3% in 2024 (data not shown).

#### LFHV-HRNZE

LFHV-HRNZE tested a total of 2483 samples between February 2021 and December 2024, of which 85.0% (n = 2099) were collected during the 2021 EVD resurgence (**Table S3**). Of those, 50.9% (n = 1069) were EDTA-blood samples and 49.1% (n = 1030) oral swabs collected in the context of community or hospital deaths. In subsequent years, the laboratory received considerably less samples ranging from 75 to 174 samples annually, all mostly EDTA-blood (**Figure 1, C-D**; **Table S3**). A differential diagnostic approach was implemented in 2022, resulting in 1480 real-time RT-PCR assays performed on 384 samples between 2022 and 2024, corresponding to approximatively 4 tests per sample, as requested by clinicians (**Figure 1, C-D**; **Table S3**).

The metadata of the samples received showed a median age of 46 years [IQR 27–67, min-max: 1-107], and both sexes similarly represented with 46.9% female (**Table S4**). The majority of samples originated from N’Zérékoré prefecture (83.8%), while the remaining were from neighboring prefectures including Beyla, Lola, Macenta, and Yomou. Students (including children in pre-school age) accounted for the largest population getting tested with 29.9%, then 25.2% domestic workers, 10.6% farmers, and 4.1% health workers of the categorized occupations (**Table S4**).

In 2021, diagnostic testing was primarily for EBOV in the context of the EVD resurgence with 14 samples (0.7%) found positive by real-time RT-PCR (**Figure 1, E**; **Table S3**). LASV real-time RT-PCR was setup in 2021 leading to the detection of 9 cases (3.9%) (**Figure 1, E**; **Table S3**). Of those 9 cases, two were identified during retrospective testing of anonymized samples as part of training or of another study, and hence not reported to health authorities (**Table S3**). In consecutive years, another 5 cases were confirmed with Lassa fever (including one case previously detected at LFHV-GKD who sought health care later at the Regional Hospital in N’Zérékoré), leading to a total of 14 cases of Lassa fever diagnosed between 2021 and 2024 at

LFHV-HRNZE, of which 12 prospectively notified to health authorities (**Figure 1, E**; **Table S3**). This corresponded to an overall RT-PCR positivity rate of 2.3% across the four years (**Table S3**). From 2022 onward, the laboratory expanded its target pathogens to include MARV, YFV and DENV. One imported DENV-positive case was detected in 2024 described in [30] (**Figure 1, E**; **Table S3**). YFV and MARV remained undetected. Laboratory TaT (i.e., sample collection to result) had a median of 0 day [IQR 0-0; min-max: 1-48]. Due to missing data, the median time from symptom onset to sample collection could not be assessed.

### Insights into Lassa fever detected between 2020-2024

Of the 30 LASV-positive diagnostics, 28 were prospective and notified to the health authorities along the 5 years while two were retrospectively detected in stored samples. Cases geolocation was strongly skewed toward prefectures hosting the two laboratories, with both Guéckédou (n = 15) and N’Zérékoré (n = 8) prefectures accounting for most of the confirmed case count (**Figures 2 and 3A; Table S5**). Yearly case detection varied with the highest number of cases in 2021 (n = 10) and in 2023 (n = 9) (**Figure 1, E; Figure 3, A**). The median age was 31 years [IQR 24-50, min max: 3-75], with males (66.7%) being more represented than females (**Table S5**). Lassa fever was diagnosed in individuals with various work categories including farmers (23.3%), domestic workers (23.3%), and students including children in pre-school age (20.0%) (**Table S5**). The symptoms recorded at presentation included fever (90%), fatigue (73%), vomiting (63%), and bleeding (60%) among others (**Figure 3, B; Table S7**). The median time from symptom onset to sample collection was 7 days [IQR 3-10, min-max: 0-22], and from onset to test result in a similar range with 7 days [IQR 3-10.5, min-max: 0-23].

**Figure 2.**
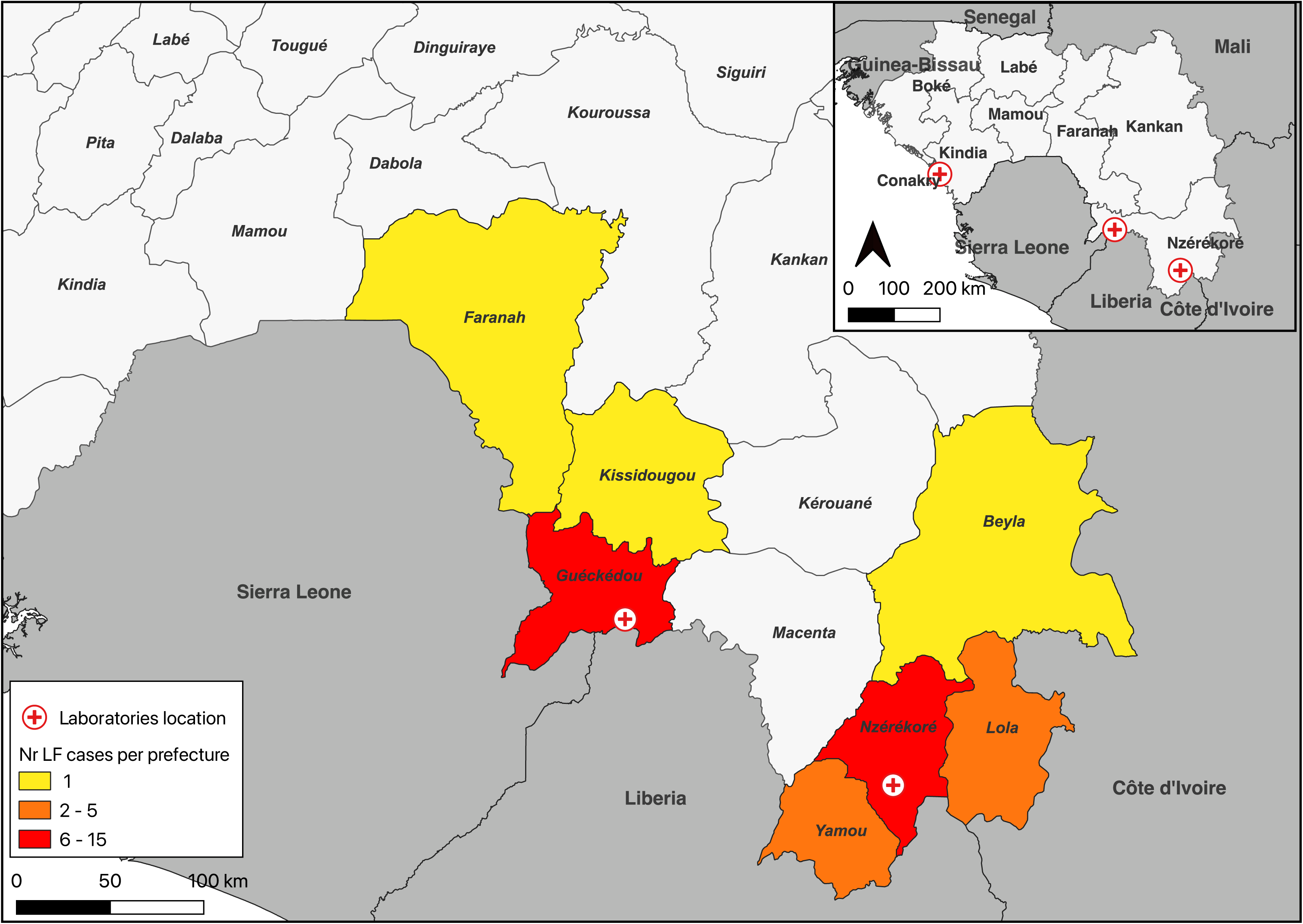
Geographical location of the labs and prefecture of Lassa fever confirmed cases. Map of Guinea showing the location of the three laboratories CRV-LFHVG (Conakry), LFHV-GKD (Guéckédou) and LFHV-HRNZE (N’Zérékoré) (inset: small square), and highlighting the prefectures of origin and the number of Lassa fever confirmed cases between 2020 and 2024. A color gradient representing the number of cases was used.

**Figure 3.**
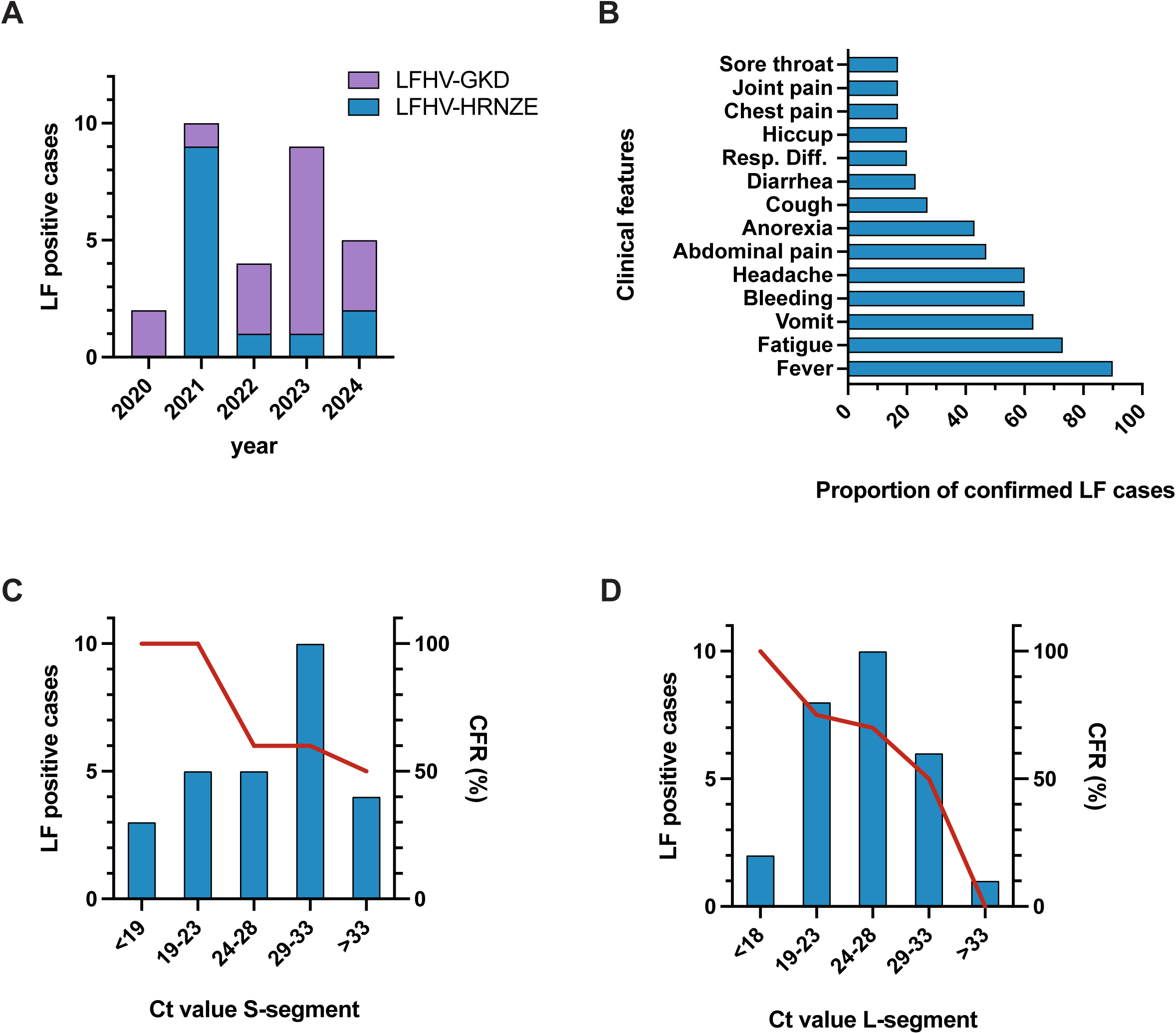
Lassa fever case counts, clinical symptoms at presentation and viral loads. (A) Number of Lassa fever positive cases detected by each laboratory per year. (B) Proportion of reported symptoms among Lassa fever confirmed cases (from Table S6). (C, D) Number of positive cases (n = 28, left axis) and their corresponding case fatality rate (CFR, right axis)) according to cycle threshold (Ct) value ranges for the S-segment (C) or L-segment (D) used as proxy for viral loads. Excluded from the graphs the cases for whom the outcome is unknown and/or one of the RT-PCR targets was over limit of detection (Ct value ≥45).

LASV RT-PCR has two targets, one for each segment of the virus, with the small (S) and large (L) segments, and the cycle threshold (Ct) values obtained can be used as a proxy for viral load, with low Ct values indicative of high viral RNA level or high viral load. LASV samples showed a median Ct value for S-segment of 30 [IQR 23-33, min-max: 14-44] and for L-segment of 26.5 [IQR 22-31, min-max: 16-44] (**Table S5**), with a strong positive correlation between the two RT-PCR targets (**Figure S1**). Case fatality rate (CFR) was higher (100% for S-segment, 80% for L-Segment) in individuals harboring Ct values ≤ 23 as compared to those with Ct values > 23 for both the two LASV targets (**Figure 3, C-D**). The overall LASV CFR was 67.9% (19/28). Median age was higher in patients who died (35 years [IQR 25-52, min-max 3-75]) compared to those who survived (25 years [IQR 22-30, min-max 17-57]). For a subset of Lassa fever cases, serological testing was performed on the initial diagnostic specimen to assess LASV serostatus (both anti-LASV IgG and IgM). Of the 22 samples tested, 31.8% were IgM-positive and 72.7% were IgG-positive (**Table S5**). No association between the LASV serostatus and or age, prefecture of origin, time from symptom onset to sample collection, and Ct values was observed (data not shown).

### Clinical management and epidemiological investigations of the Lassa fever cases

Following laboratory confirmation, all patients were immediately isolated in the treatment centers (CTEpi). A total of 10 cases (35.7%) were treated with ribavirin (**Table S7**), while treatment could not be provided to others due to (i) either limited drug availability in country, or (ii) clinical contraindications including pregnancy or severe disease with rapid death. Otherwise, clinical care of Lassa fever cases was supportive. Viremia follow-up appeared limited and conducted at the clinician’s discretion.

Among the 10 female cases, three (F1, F2 and F3) were pregnant. Two of them, F2 and F3, died presenting high viremia as inferred from their Ct values (Ct values of S- and L-segments of 18 and 18 for F2, and of 18 and 19 for F3, respectively). F1 survived and had Ct values for S- and L-segment of 34 and 31, respectively. All three presented at the hospital within 3 to 8 days from disease onset and displayed bleeding symptoms (no other information available). Due to contraindications, none of them received ribavirin treatment. Two women (F1 and F2) experienced miscarriages following hospitalization, and one (F3) delivered an alive infant who died a few days after birth.

Of the 28 Lassa fever cases prospectively detected, epidemiological investigations pointed towards a transmission from the natural rodent reservoirs, and no secondary transmissions were detected. Contacts of the primary case were monitored for up to 21 days post-exposure, and tested by RT-PCR only if symptoms that meet the suspected VHFs case definition arose. Of the 28 cases, two reported prior travel to Sierra Leone and Liberia before symptom onset. While no others reported trips prior to onset, short-duration travels could not be excluded from the epidemiological investigations.

At the request of the MoH, ecological and environmental investigations were conducted in the villages of origin of selected Lassa fever cases as rodents were identified as the likely source of infection. Seven investigations were carried out between 2022 and 2024, during which rodents from the household, neighboring houses and or the geographical area were trapped.

Rodent excreta and blood samples were collected and tested for LASV at LFHV-GKD and LFHV-HRNZE. LASV was detected in rodent samples in only one investigation site from 2022 in the Kassadou sub-prefecture of the Guéckédou prefecture.

## Discussion

Before EVD outbreak of 2014-2016, routine laboratory-based surveillance of high consequence pathogens was lacking in the forested region of Guinea. We show here major improvement in such surveillance at tertiary health institutions level with the establishment and strengthening of two laboratories offering routine VHF diagnostics without charge.

Setup in the endemic region, LFHV-GKD and -HRNZE lab preparedness was achieved through a long-term program enabling the testing of five high-priority viruses in remote, decentralized, and resource-limited regions where diagnostic capacity was previously absent [31]. This contributed to substantial support to Guinea’s health system leading to the timely identification of several VHF outbreaks or events, including the previously reported EVD, Marburg virus disease, dengue fever and Lassa fever [13,26,29,30], and hereby the report on one additional yellow fever case and 30 Lassa fever cases identification.

As in 2021, the effective local readiness of LFHV-GKD during the EVD resurgence highlighted the critical need to preserve and further decentralize laboratory capacity in regions at elevated risk for VHFs, which is reiterated with the new findings here [32,33]. Expanding the network of surveillance to an additional laboratory in N’Zérékoré enabled the detection of 13 LASV positive cases which otherwise may not have been detected.

The availability of a diverse diagnostic portfolio facilitated effective case detection, to the benefit of the population. Access to complementary molecular diagnostics is crucial for VHFs diagnosis due to the nonspecific nature of their symptoms, which complicate etiology confirmation and patient management. Real-time RT-PCR platforms and assays, as used here, offer such versatility to flexibly and broadly investigate relevant pathogens at family or genus level (e.g., filoviruses). This approach requires long-term training, mentoring and precision work to handle such setup with safe inactivation of samples, genetic material extraction, RT-PCR setup, and also to mitigate the risk of contamination, all those requiring adequate and stringent quality assurance processes in place. The use of point-of-care diagnostic tests is often advocated to simplify ease of use, broad access to non-specialized staff or decrease TAT, though such methodology highly limit versatility and resourcefulness to respond to future threats and may limit detection at virus species level [34–36]. TAT were generally short in both laboratories demonstrating availability of reagents, and effective transport systems for sample dispatch in place, as well as optimal sample processing time to results which is highly relevant for quick public health intervention in the context of VHFs.

Despite limited and constrained infrastructure, the two laboratories have processed a substantial number of samples and provided a wide range of diagnostic testing free of charge, including LASV. Previous seroprevalence studies reporting high LASV exposure in this region suggesting virus exposure with yet a lack of acute cases being detected [12,16,17]. Here, we show that acute Lassa fever cases occur with 30 confirmed between 2020 and 2024 across both laboratories, with a positive detection rate ranging between 1.4% to 2.3%. Through the collaborative network of the three laboratories for VHFs surveillance in country, samples were further sequenced at the national reference lab (CRV-LFHVG), providing new knowledge about LASV genomic diversity in human cases in Guinea [37].

Lassa fever CFR of 67.9% observed here is higher than those previously reported from Guinea of 18% [12] and Liberia of 40% [38], but similar to the 69% from Sierra Leone [39]. Of course, a bias exists towards detection of febrile individuals seeking for medical care whom may already be advanced in disease progression. Less than 40% of the Lassa fever patients received ribavirin, largely reflecting limited drug availability and the occurrence of early deaths before treatment initiation. The median time between symptom onset and sample collection or test result was 7 days, consistent with findings from a previous report in Nigeria [40]. Presentation more than 7 days after symptom onset is considered delayed, as this falls beyond the optimal window for early patient management, including timely supportive care and initiation of treatment [24]. Improving awareness on Lassa fever, particularly in rural communities, and strengthening first line health services (such as primary health centers) to support earlier and faster case detection could facilitate in the future timelier diagnosis and prompt case management. This is especially critical for pregnant women, in whom bleeding is often misinterpreted, leading to delayed diagnosis and substantial adverse consequences for both maternal and child health, as reported also here [41].

Substantial limitations exist in our study, specifically regarding the investigation on Lassa fever cases. Clinical data were not systematically collected, and basic clinical management information was lacking or incomplete, limiting the validity and generalizability of the clinical findings. As reported in other studies [40], higher CFR is seen in patients with high viremia or Ct values ≤ 23 than in those with Ct values > 23; however, the limited and non-standardized data collection, as in an observational study, prevented adjustment for other factors which may influence viral load detection and or mortality. Most of the Lassa fever cases, approximately 75%, originated from Guéckédou and N’Zérékoré prefectures, where the laboratories are located, introducing a geographical bias in the descriptive analysis. Nonetheless, these observations underscore the critical role of laboratory presence locally in reinforcing health system capacity. None of the Lassa fever cases identified through the surveillance activities in the two laboratories originated from the prefecture of Macenta, although samples from this area were received for testing. Given that high seroprevalence for LASV was reported in the area [42], further investigation is warranted to determine whether the observation here reflects limitation in the surveillance coverage or differences in virus circulation. Generally, the expansion of diagnostic and surveillance capacity to additional regions in country may help improve case detection, not only for Lassa fever but for VHFs.

Our work emphasizes also the importance of sustainable funding for effective preparedness against VHFs and emerging viral diseases. The Global Health Protection Programme (GHPP) of the German Ministry of Health was key in long-term development of theses capacities and public health findings. However, such capacity strengthening support funds are now becoming increasingly limited globally which may jeopardize the years long built country assets. Sustaining this capacity remains highly challenging, particularly within national health systems in low- and middle-income countries which are already fragilized and where laboratory and surveillance activities often rely on the WHO and other external support, which are now facing major financial constraints, threatening health systems and global health more broadly.

Post-EVD outbreak (2014-2016), Guinea has improved its knowledge, attitudes, and practices toward VHFs, though gaps persist, particularly in rural healthcare facilities and on Lassa fever management [43]. Our study emphasizes the need for investment in laboratory infrastructure and personnel training in endemic areas. Strengthening local diagnostic capacity not only improves disease surveillance but also enhances overall health system resilience, with evidence from other African countries, consistent with our findings, showing that local laboratory presence enables earlier case detection and timely patient management [32]. These results underscore the critical importance of maintaining laboratory capacity in forested areas at high risk of spillover events. Future research should assess the long-term impact of enhanced laboratory capacity on early detection and mortality and explore novel diagnostic tools, such as rapid antigen tests, to further accelerate case identification. We also advocate with this study for increased awareness of Lassa fever in Guinea, a disease often neglected due to its low incidence in the country.

## Ethical approval

This descriptive research, using anonymized diagnostic surveillance data, has been approved by the National Ethics Committee of Guinea (CNERS) under the number 197/CNERS/25 and 189/CNERS/24.

## Consent for publication

Not applicable.

## Declaration of generative AI and AI-assisted technologies in the writing process

During the preparation of this work the author(s) used ChatGPT / free version to edit some sentences. After using this tool/service, the author(s) reviewed and edited the content as needed and take(s) full responsibility for the content of the publication.

## Contributions

Conceived and designed the study: F.R.K., Y.S., S.G., S.B., N.F.M., S.Du., G.A. Collected data and/or Performed laboratory diagnostics: F.R.K., Y.S., S.L.M., K.I., J.H., H.S., Kar.K., T.E.M., F.M.T., M.K., M.D.B., S.K., B.S., M.T., Kab.K., M.H., S.R., C.v.G., G.A. Project implementation: F.R.K., Y.S., S.L.M., K.I., J.H., H.S., M.H., S.R., C.v.G., B.B.Z., C.J., A.T., L.O., A.L., P.F., A.A.K., S.Di., M.F.J.M.K., M.P., S.G., K.M., Kab.K., S.H.G., S.B., N.F.M., S.Du., G.A.

Funding acquisition: A.L., P.F., M.P., S.G., S.B., N.F.M., S.Du.

Wrote the manuscript: F.R.K., Y.S., H.S., S.G., N.F.M, S.Du., G.A.

Edited the manuscript: all authors.

All authors read and approved the content of the manuscript.

## Data Availability

All data produced in the present study are available upon reasonable request to the authors

## Acknowledgments

The authors thank the Ministry of Health of the Republic of Guinea, and all healthcare workers involved in the surveillance activities. We would like to acknowledge Prof. Miles Carroll and Dr. Joseph Akoi Boré for their contributions to the initial establishment and setup of the laboratory in Guéckédou. We acknowledge Ndapewa Ithete for her thorough review of the manuscript.

## Supplementary file

**Table S1.**
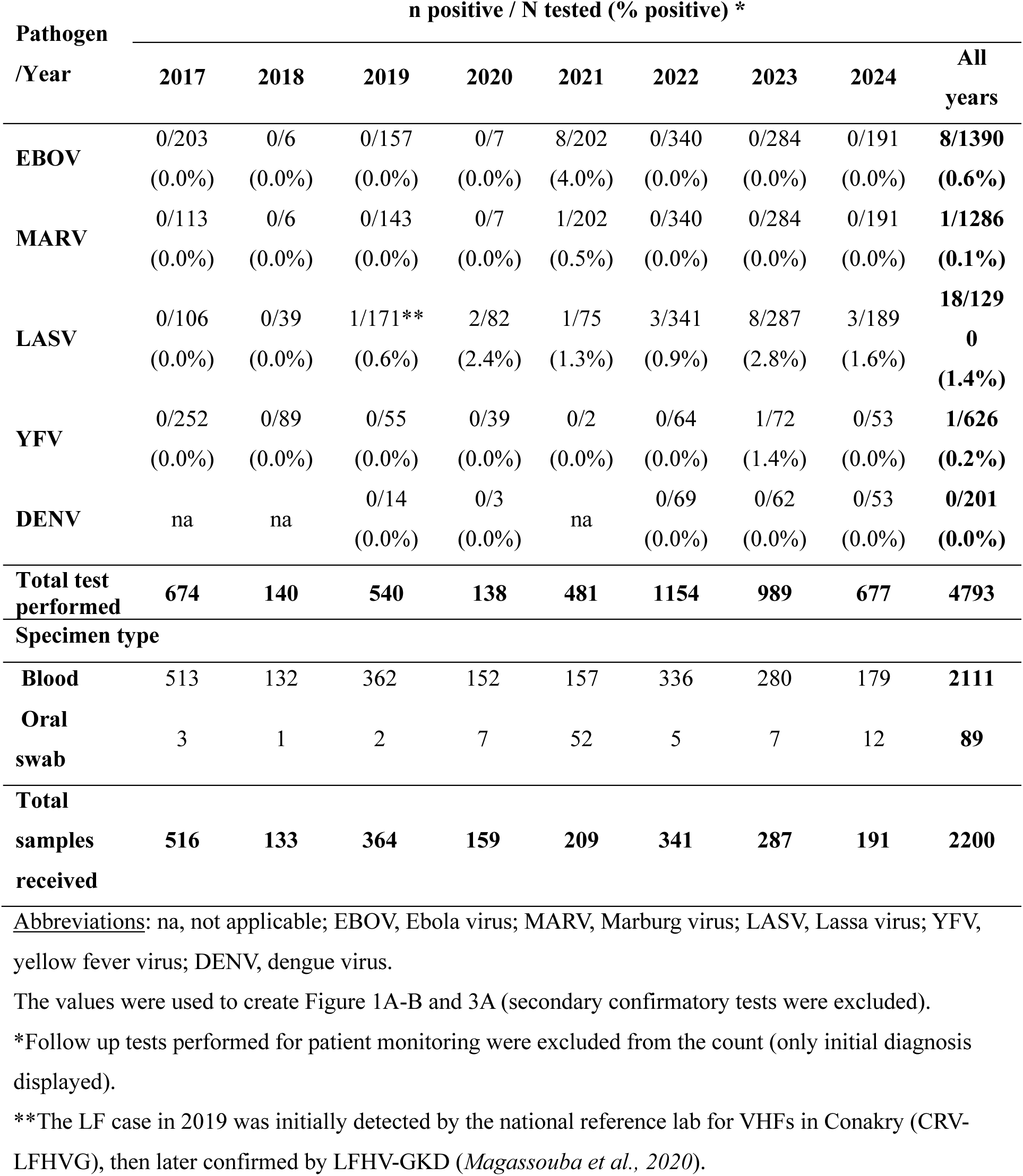
VHFs diagnostics testing performed at LFHV-GKD, 2017-2024.

| **Pathogen**<br>**/Year** | **n positive / N tested (% positive) \*** |  |  |  |  |  |  |  | **All**<br>**years** |
| --- | --- | --- | --- | --- | --- | --- | --- | --- | --- |
|  | **2017** | **2018** | **2019** | **2020** | **2021** | **2022** | **2023** | **2024** |  |
| **EBOV** | 0/203<br>(0.0%) | 0/6<br>(0.0%) | 0/157<br>(0.0%) | 0/7<br>(0.0%) | 8/202<br>(4.0%) | 0/340<br>(0.0%) | 0/284<br>(0.0%) | 0/191<br>(0.0%) | **8/1390**<br>**(0.6%)** |
| **MARV** | 0/113<br>(0.0%) | 0/6<br>(0.0%) | 0/143<br>(0.0%) | 0/7<br>(0.0%) | 1/202<br>(0.5%) | 0/340<br>(0.0%) | 0/284<br>(0.0%) | 0/191<br>(0.0%) | **1/1286**<br>**(0.1%)** |
| **LASV** | 0/106<br>(0.0%) | 0/39<br>(0.0%) | 1/171**<br>(0.6%) | 2/82<br>(2.4%) | 1/75<br>(1.3%) | 3/341<br>(0.9%) | 8/287<br>(2.8%) | 3/189<br>(1.6%) | **18/1290**<br>**(1.4%)** |
| **YFV** | 0/252<br>(0.0%) | 0/89<br>(0.0%) | 0/55<br>(0.0%) | 0/39<br>(0.0%) | 0/2<br>(0.0%) | 0/64<br>(0.0%) | 1/72<br>(1.4%) | 0/53<br>(0.0%) | **1/626**<br>**(0.2%)** |
| **DENV** | na | na | 0/14<br>(0.0%) | 0/3<br>(0.0%) | na | 0/69<br>(0.0%) | 0/62<br>(0.0%) | 0/53<br>(0.0%) | **0/201**<br>**(0.0%)** |
| **Total test performed** | **674** | **140** | **540** | **138** | **481** | **1154** | **989** | **677** | **4793** |
| **Specimen type** |  |  |  |  |  |  |  |  |  |
| **Blood** | 513 | 132 | 362 | 152 | 157 | 336 | 280 | 179 | **2111** |
| **Oral swab** | 3 | 1 | 2 | 7 | 52 | 5 | 7 | 12 | **89** |
| **Total samples received** | **516** | **133** | **364** | **159** | **209** | **341** | **287** | **191** | **2200** |

Abbreviations: na, not applicable; EBOV, Ebola virus; MARV, Marburg virus; LASV, Lassa virus; YFV, yellow fever virus; DENV, dengue virus.
The values were used to create Figure 1A-B and 3A (secondary confirmatory tests were excluded).
\*Follow up tests performed for patient monitoring were excluded from the count (only initial diagnosis displayed).
\*\*The LF case in 2019 was initially detected by the national reference lab for VHFs in Conakry (CRV-LFHVG), then later confirmed by LFHV-GKD (*Magassouba et al., 2020*).

**Table S2.** Sociodemographic characteristics of VHFs suspected cases tested at LFHV-GKD, 2017-2024.

| Characteristics | Frequency (percentage) |
| --- | --- |
| **Age (years)\*\*** |  |
| Median (IQR), min-max: 35 (24-50), 1-95 |  |
| **Sex** |  |
| Female | 1252/2200 (56.5%) |
| Male | 958/2200 (43.5%) |
| **Occupation** |  |
| Domestic worker | 762/2200 (34.7%) |
| Farmer | 337/2200 (15.3%) |
| Health worker | 75/2200 (3.4%) |
| Other | 603/2200 (27.4%) |
| Student* | 357/2200 (16.2%) |
| Unknown | 66/2200 (3.0%) |
| **Prefecture of residence\*\*** |  |
| Guéckédou | 2099/2192 (95.8%) |
| N'Zérékoré | 31/2192 (1.4%) |
| Other | 34/2192 (1.5%) |
| Yomou | 28/2192 (1.3%) |

Abbreviations: IQR, Interquartile range.
\*Including children in pre-school age (e.g. 3 years old).
\*\*Total number differs from total, missing data.

**Table S3.** VHFs diagnostics testing performed at HRNZE, 2021-2024.

| **Pathogen /Year** | **n positive / N tested (% positive) \*** |  |  |  |  |
| --- | --- | --- | --- | --- | --- |
|  | **2021** | **2022** | **2023** | **2024** | **All years** |
| **EBOV** | 14/1972**<br>(0.7%) | 0/70<br>(0.0%) | 0/172<br>(0.0%) | 0/133<br>(0.0%) | **14/2347**<br>**(0.6%)** |
| **MARV** | 0/3<br>(0.0%) | 0/70<br>(0.0%) | 0/172<br>(0.0%) | 0/133<br>(0.0%) | **0/378**<br>**(0.0%)** |
| **LASV\*\*\*** | 9/231<br>(3.9%) | 2/71<br>(2.8%) | 1/174<br>(0.6%) | 2/135<br>(1.5%) | **14/611**<br>**(2.3%)** |
| **YFV** | na | 0/12<br>(0.0%) | 0/91<br>(0.0%) | 0/83<br>(0.0%) | **0/186**<br>**(0.0%)** |
| **DENV** | na | 0/28<br>(0.0%) | 0/53<br>(0.0%) | 1/83<br>(1.2%) | **1/164**<br>**(0.6%)** |
| **Total test performed** | **2206** | **251** | **662** | **567** | **3686** |
| **Specimen type** |  |  |  |  |  |
| **Blood** | 1069 | 73 | 174 | 130 | **1466** |
| **Oral swab** | 1030 | 2 | 0 | 5 | **1037** |
| **Total samples received** | **2099** | **75** | **174** | **135** | **2483** |

Abbreviations: na, not applicable; EBOV, Ebola virus; MARV, Marburg virus; LASV, Lassa virus; YFV, yellow fever virus; DENV, dengue virus.
The values in the table were used to create Figure 1C-D and 3A (secondary confirmatory tests were excluded).
\*Follow up tests performed for patient monitoring were excluded from the count (only initial diagnosis displayed).
\*\*Includes testing by real time RT-PCR and RDTs; six of the EBOV positive cases had prior positive results at LFHV-GKD (confirmatory testing).
\*\*\*In 2021, of the 9 LF cases, 7 were detected through prospective testing, while 2 were from retrospective investigation; in 2022 one LF case tested at HRNZE corresponds to a LF case previously detected at LFHV-GKD who sought health care later at the Regional Hospital in N'Zérékoré.

**Table S4.**
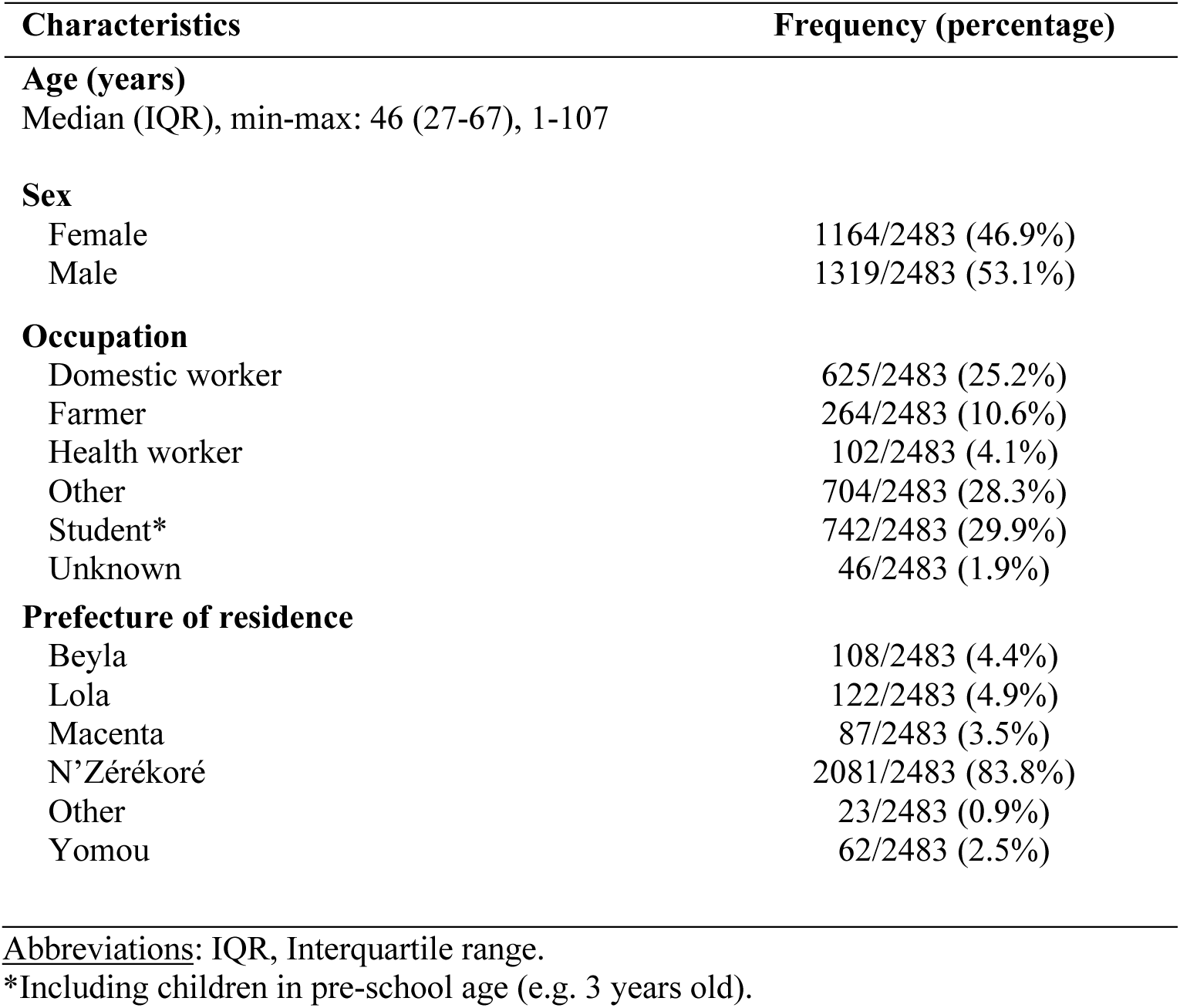
Sociodemographic characteristics of VHFs suspected cases tested at HRNZE, 2021-2024.

| **Characteristics** | **Frequency (percentage)** |
| --- | --- |
| **Age (years)** |  |
| Median (IQR), min-max: 46 (27-67), 1-107 |  |
| **Sex** |  |
| Female | 1164/2483 (46.9%) |
| Male | 1319/2483 (53.1%) |
| **Occupation** |  |
| Domestic worker | 625/2483 (25.2%) |
| Farmer | 264/2483 (10.6%) |
| Health worker | 102/2483 (4.1%) |
| Other | 704/2483 (28.3%) |
| Student* | 742/2483 (29.9%) |
| Unknown | 46/2483 (1.9%) |
| **Prefecture of residence** |  |
| Beyla | 108/2483 (4.4%) |
| Lola | 122/2483 (4.9%) |
| Macenta | 87/2483 (3.5%) |
| N'Zérékoré | 2081/2483 (83.8%) |
| Other | 23/2483 (0.9%) |
| Yomou | 62/2483 (2.5%) |

Abbreviations: IQR, Interquartile range.
\*Including children in pre-school age (e.g. 3 years old).

**Table S5.** Sociodemographic characteristics of Lassa fever cases, Guinea 2020-2024.

| **Characteristics** | **Frequency (percentage)** |
| --- | --- |
| **Age (years)** |  |
| Median (IQR), min-max = 31 (24-50), 3-75 |  |
| <25 years | 8/30 (26.7%) |
| 25-30 years | 7/30 (23.3%) |
| 31-49 years | 7/30 (23.33%) |
| >49 years | 8/30 (26.7%) |
| **Sex** |  |
| Female | 10/30 (33.3%) |
| Male | 20/30 (66.7%) |
| **Occupation** |  |
| Domestic worker | 7/30 (23.3%) |
| Farmer | 7/30 (23.3%) |
| Health worker | 2/30 (6.7%) |
| Other | 8/30 (26.7%) |
| Student* | 6/30 (20.0%) |
| **Prefecture of residence** |  |
| Beyla | 1/30 (3.3%) |
| Faranah | 1/30 (3.3%) |
| Guéckédou | 15/30 (50.0%) |
| Kissidougou | 1/30 (3.3%) |
| Lola | 2/30 (6.7%) |
| N'Zérékoré | 8/30 (26.7%) |
| Yomou | 2/30 (6.7%) |
| **Year** |  |
| 2020 | 2/30 (6.7%) |
| 2021** | 10/30 (33.3%) |
| 2022 | 4/30 (13.3%) |
| 2023 | 9/30 (30.0%) |
| 2024 | 5/30 (16.7%) |

Abbreviations: IQR, Interquartile range.
The values were used to create Figure 2.
\*Including children in pre-school age (e.g. 3 years old)
\*\*Including two samples from retrospective testing, not notified to MoH

**Table S6.** Symptoms at presentation among Lassa fever cases, Guinea 2020-2024.

| Symptom* | Frequency (percentage) |
| --- | --- |
| Fever | 27/30 (90%) |
| Fatigue | 22/30 (73%) |
| Vomit | 19/30 (63%) |
| Bleeding | 18/30 (60%) |
| Headache | 18/30 (60%) |
| Abdominal pain | 14/30 (47%) |
| Anorexia | 13/30 (43%) |
| Cough | 8/30 (27%) |
| Diarrhea | 7/30 (23%) |
| Resp. Diff. | 6/30 (20%) |
| Hiccup | 6/30 (20%) |
| Chest pain | 5/30 (17%) |
| Joint pain | 5/30 (17%) |
| Sore throat | 5/30 (17%) |

\*As recorded on laboratory request form accompanying the sample. The values were used to create Figure 3B.

**Table S7.**
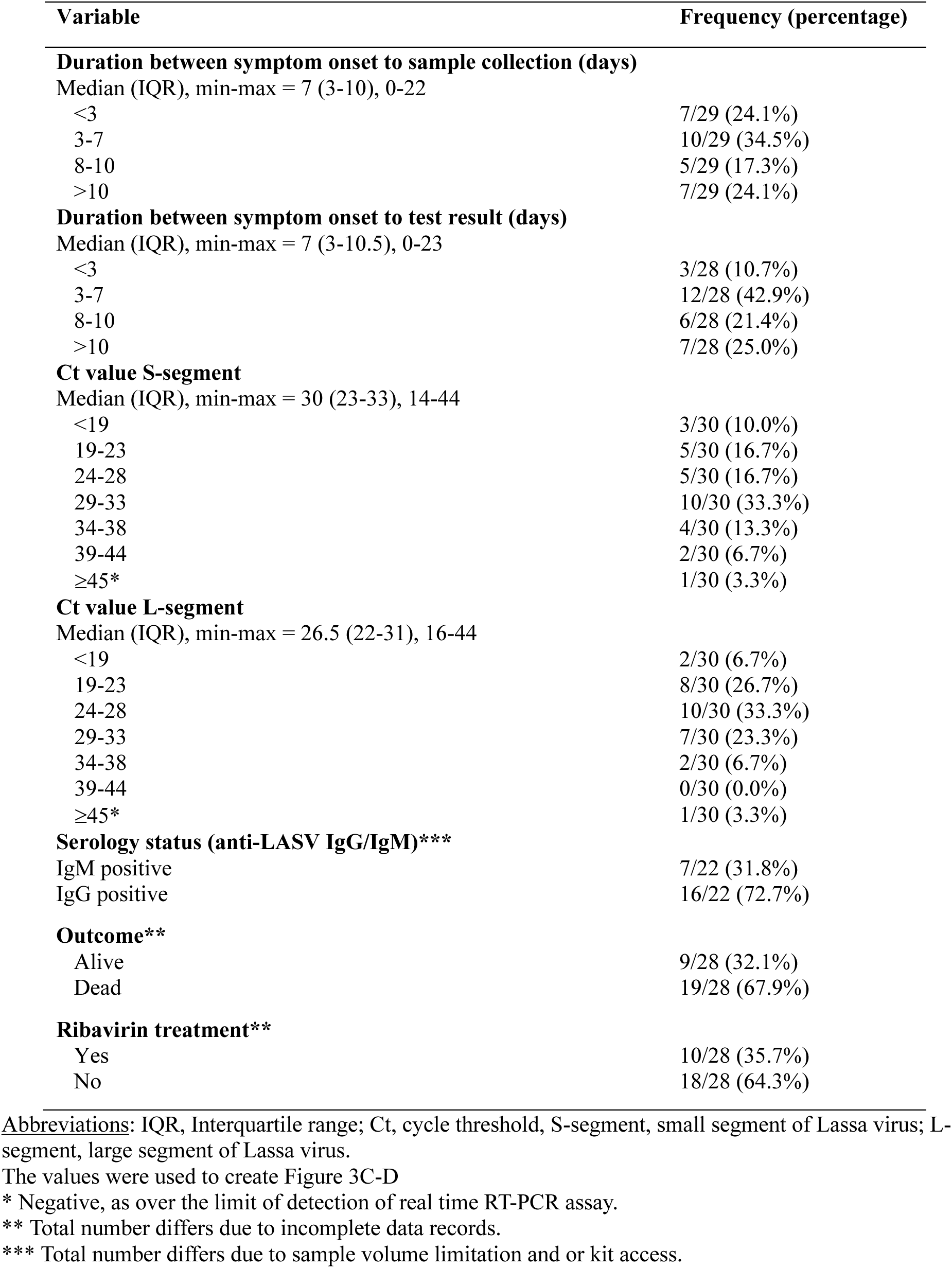
Outcome, laboratory findings and ribavirin treatment among Lassa fever cases, Guinea 2020-2024.

**Figure S1.**
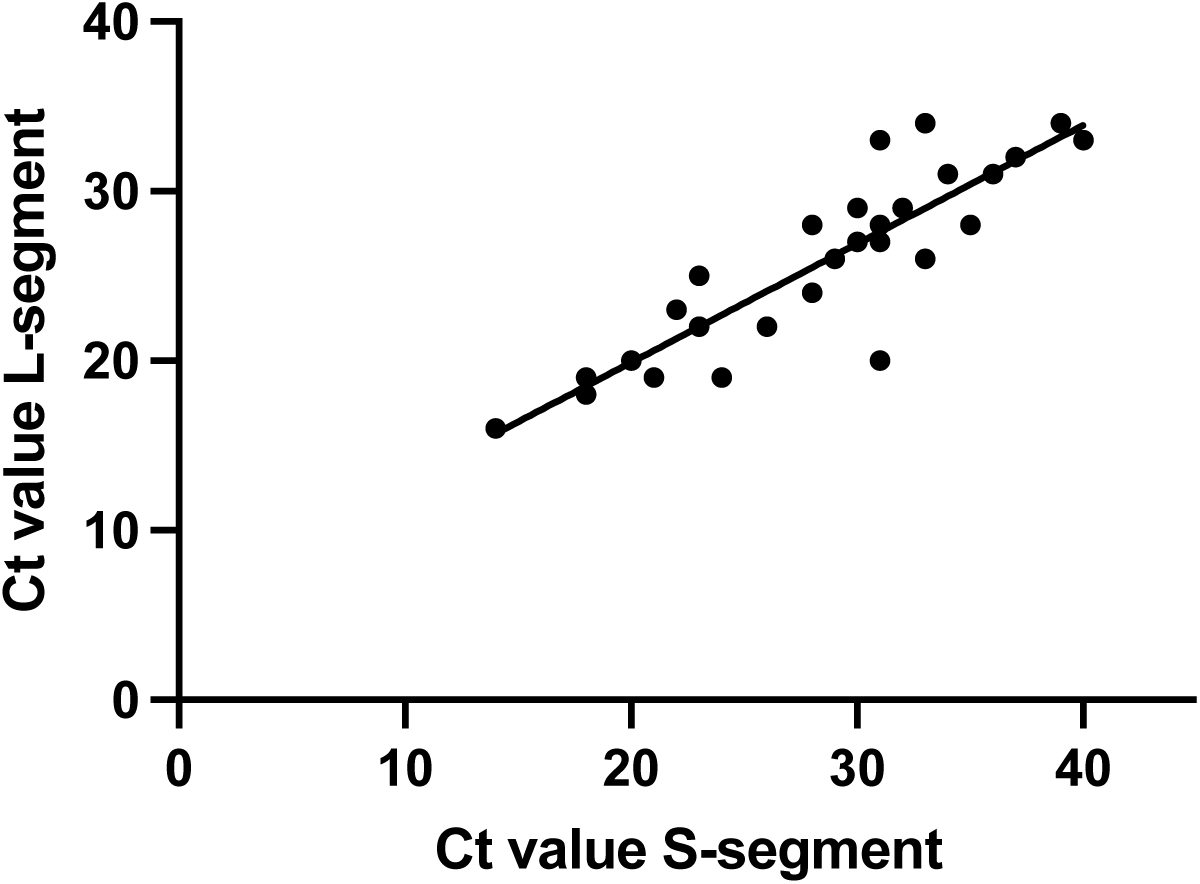
Correlation between Ct values for S-segment and L-segment among Lassa fever cases, Guinea 2020-2024. Scatter plot of real-time RT–PCR Ct values for S-segment (x-axis) and L-segment (y-axis) from LASV–positive samples (n=30). Each point represents a sample (first test only included). A strong positive association was observed (R²=0.7708; p<0.0001). The fitted regression line is shown (Y=0·6999×X+5·900).

